# Skin imprints to provide noninvasive metabolic profiling of COVID-19 patients

**DOI:** 10.1101/2021.04.17.21255518

**Authors:** Jeany Delafiori, Rinaldo Focaccia Siciliano, Arthur Noin de Oliveira, José Carlos Nicolau, Geovana Manzan Sales, Talia Falcão Dalçóquio, Estela Natacha Brandt Busanello, Adriana Eguti, Diogo Noin de Oliveira, Adriadne Justi Bertolin, Luiz Augusto dos Santos, Rocío Salsoso, Fabiana G Marcondes-Braga, Nelson Durán, Maurício Wesley Perroud, Ester Cerdeira Sabino, Leonardo Oliveira Reis, Wagner José Fávaro, Rodrigo Ramos Catharino

## Abstract

As the current COVID-19 pandemic progresses, more symptoms and signals related to how the disease manifests in the human body arise in the literature. Skin lesions and coagulopathies may be confounding factors on routine care and patient management. We analyzed the metabolic and lipidic profile of the skin from COVID-19 patients using imprints in silica plates as a non-invasive alternative, in order to better understand the biochemical disturbances caused by SARS-CoV-2 in the skin. One hundred and one patients (64 COVID-19 positive patients and 37 control patients) were enrolled in this cross-sectional study from April 2020 to June 2020 during the first wave of COVID-19 in São Paulo, Brazil. Fourteen biomarkers were identified related to COVID-19 infection (7 increased and 7 decreased in COVID-19 patients). Remarkably, oleamide has shown promising performance, providing 79.0% of sensitivity on a receiver operating characteristic curve model. Species related to coagulation and immune system maintenance such as phosphatidylserines were decreased in COVID-19 patients; on the other hand, cytokine storm and immunomodulation may be affected by molecules increased in the COVID-19 group, particularly primary fatty acid amides and N-acylethanolamines, which are part of the endocannabinoid system. Our results show that skin imprints may be a useful, noninvasive strategy for COVID-19 screening, by electing a pool of biomarkers with diagnostic potential.

## INTRODUCTION

Over a year after the global pandemic was announced, the use of non-pharmaceutical interventions, such as diagnostic methods, use of masks, hygiene and social distancing remain essential to prevent viral transmission. Meanwhile, research and initiatives to obtain more information regarding COVID-19 pathophysiological mechanisms are still ongoing^1^. Many efforts to understand the pathogenic mechanisms of SARS-CoV-2 infection are based on human metabolic evaluation, which has been showing the involvement of an intense immune response, as well as dysfunctional coagulation and inflammation patterns^2^. Several studies employing “omics” approaches indicate altered expression of proteins and disrupted lipid metabolism^3,4^, suggesting the applicability of metabolite investigation at proteomic^4^, metabolomic^4–10^ and lipidomic^3,9–13^ levels. Most contributions employed human samples with established clinical importance, such as plasma^5,8–10^, serum^3,4,13^, and saliva^7^. Nevertheless, novel approaches such as nasal excretion^11^, exhaled breath^6^, and sebum^12^ are under investigation as potential sources of biomarkers that discriminate COVID-19 for diagnosis, and especially for prognosis purposes. Spick et al. (2021) demonstrated that triacylglycerides alterations in sebum lipidome have the potential to be used as a screening test for COVID-19 ^12^. Consequently, the broad range of disrupted metabolites observed in such diverse sample types highlights the comprehensive spectrum of the infection manifestations.

The distribution of SARS-CoV-2 cell entry receptor assists in characterizing COVID-19’s diversity of symptoms that portray multi-organ involvement^14^. ACE2 (angiotensin-converting enzyme 2) was identified as a crucial receptor for SARS-CoV-2 binding and cell entry^14,15^. High levels of ACE2 expression are present in different cell types of multiple organs, such as oral cavity, lungs, gastrointestinal tract, and even keratinocytes in the skin^14,15^. When intact, skin is not considered a gateway for the virus; thus, skin contacts have not been determined as a SARS-CoV-2 transmission route. Conversely, skin lesions have been reported in several cohorts as an underlying manifestation of COVID-19, including in asymptomatic cases^16,17^. The most common observed skin manifestations were erythematous-violaceous lesions, chilblain-like acral lesions, erythematous rash, urticarial rash, papulovesicular exanthem, and purpuric lesions that might be associated with the overall inflammatory state, and altered thrombotic and coagulation patterns due to COVID-19^14,16,17^.

In clinical practice, skin manifestations are rare when compared to other symptoms, and are usually not considered as the first signs of COVID-19^16^. However, sampling metabolites found on normal skin surfaces may offer a powerful resource for the development of rapid and noninvasive diagnostic methods that bear minimal discomfort for the patients. Previous literature brings contributions where approaches using skin imprint in silica plates coupled with mass spectrometry were used to provide metabolic insights on leprosy^18^ and cystic fibrosis^19^. In addition to the potential for diagnosis, information on metabolic alterations present on normal skin surfaces during SARS-CoV-2 infection are still incipient and may help the scientific community to better understand the pathophysiology of COVID-19^12^. With this aim, we enrolled a cohort of 101 subjects divided into positive and negative for COVID-19 and used a clean, simple, and rapid method for skin imprint samples collection using silica plates to assess biomarkers associated with COVID-19 pathology using mass spectrometry and untargeted metabolomics.

## EXPERIMENTAL SECTION

### Study design

Patients were recruited at Hospital das Clínicas, Faculdade de Medicina, Universidade de São Paulo (HCFMUSP) localized in São Paulo city in Brazil from April to June 2020. The study was conducted according to principles expressed in the Declaration of Helsinki. Ethical approval was given by Hospital das Clínicas da Faculdade de Medicina da Universidade de São Paulo - HCFMUSP ethics committee (CAAE 32077020.6.0000.0005), and a signed consent form was collected from participants before enrolment.

Adult patients (aged 18 and over), with one or more clinical symptoms for SARS-CoV-2 infection in the last 7 days (fever, dry cough, dyspnea, and/or malaise) were eligible, recruited and tested for SARS-CoV-2 infection using nasopharyngeal samples for gold-standard RT-PCR following local protocol based on Charité and WHO recommendations^20^. Following these procedures, enrolled participants (n = 101) were separated into positive (COVID-19, CV = 64) and negative (Control, CT = 37) groups and underwent protocol for skin imprint sample collection (see section below). A schematic representation of study design and patient summarized data is available in Figure 1.

**Figure 1.**
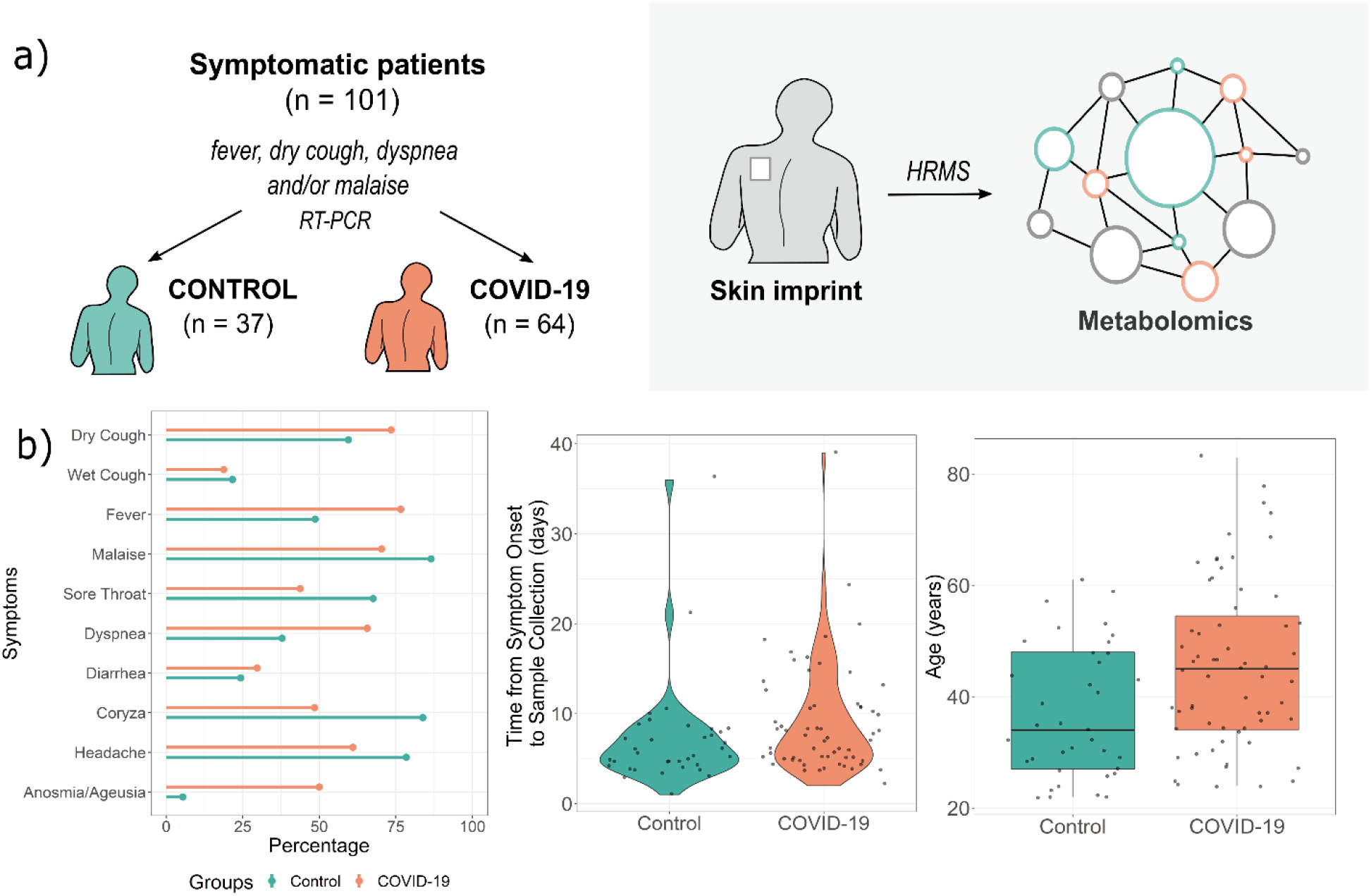
Study design (a) and patient symptoms distributions and age characteristics (b). HRMS – high-resolution mass spectrometry, RT-PCR – reverse transcriptase-polymerase chain reaction.

### Sample collection and preparation

Plates of silica gel 60 G (Merck, Darmstadt, Germany) with an area of 1 cm^2^ were used for skin imprint collection in a similar approach described for leprosy and cystic fibrosis screening^18,19^. The silica side of the plate was carefully positioned on patients’ left-back and gently pressed against the skin for 1 minute. No cleaning procedures were performed on the skin before sample collection. After the collection time, silica plates were stored in plastic tubes at −30°C until analysis. On the moment of analysis, silica plates were washed and covered with 500 µL of a Methanol:H2O (1:1) solution, agitated in vortex for 30 s, and left to decant for 5 minutes. A 200-µL aliquot of the supernatant was collected and ionized with formic acid (0.1% final concentration) before analysis.

### High-Resolution Mass Spectrometry Analysis

Groups were randomized intra- and inter-daily and directly infused into a high-resolution ESI-Q-Exactive Orbitrap (Thermo Scientific, Germany) with 140,000 FWHM resolution on positive ion mode. Spectral data were acquired following the parameters: *m/z* range 150 - 1700, 10 acquisitions per sample, 40 scans per acquisition, flow injection rate 10 µl.min^−1^, capillary temperature 320°C, spray voltage 3.70 kV, nitrogen sheath gas 8 (arbitrary units), aux gas heater temperature 30°C, RF-lens 50, AGC target 10^6^.

### Statistical analysis and biomarker selection

Patient characteristics were expressed as the count followed by frequency (percentage) for categorical variables. Groups – infected and non-infected patients – were compared using Chi-Squared independent test or Fisher’s exact test, when applicable. For continuous variables (age, onset of symptoms) normality of data distribution was tested using Shapiro-Wilk test, and the median [interquartile range (IQR)] was attributed and analyzed using the Mann-Whitney hypothesis test. P-value was considered significant if < 0.05 two-tailed. A peak intensity table was extracted from acquired spectral data; a median value was attributed for each feature considering 10 replicates acquired per sample. Data was posteriorly normalized (quantile) and transformed (logarithm) before multivariate statistical analysis using MetaboAnalyst 5.0 online software (www.metaboanalyst.ca)^21^. A PLS-DA (Partial Least Square – Discriminant Analysis) was applied to the full dataset, aiming to elect a list of discriminant *m/z* features for molecule annotation. Using the VIP score list > as reference, we consulted METLIN (Scripps Center for Metabolomics, La Jolla, CA, USA – www.metlin.scripps.edu) and LIPIDMAPS (University of California, San Diego, CA, USA - www.lipidmaps.org) databases for molecule annotation with mass accuracy < 5 ppm. The model with the annotated biomarkers was evaluated through permutation tests (n = 1000). Fold-change values and p-value significances were attributed to each identified biomarker using volcano plot (t-test) resource. Markers were used to project a prediction models using a receiver operating characteristic curve (ROC) resource with a Linear SVM algorithm from MetaboAnalyst. Permutation tests (n = 1000) were employed to validate the refined model. R coding was used to draw the graphics.

## RESULTS

### Patient characteristics

The cohort described in this study recruited 101 patients from April 2020 to June 2020 that arrived at Clinics Hospital of the University of São Paulo reporting COVID-19 symptoms, from which 64 (63.4%) tested positive for SARS-COV-2. This population was female in its majority, on both COVID-19 and Control groups (59.4% and 78.4%, respectively), with a general median age of 45 (IQR 22) and 34 (IQR 21) years old (COVID-19 and Control, respectively). Time from the onset of symptoms to sample collection was longer in COVID-19 group (median 8, IQR 6) and differed from Control (median 5.5, IQR 4) with a p-value of 0.031. The most common reported symptoms in COVID-19 patients were fever (76.6%), dry cough (73.4), malaise (70.3%), and dyspnea (65.6%); however, significant difference (p-value < 0.05) between groups were observed only for fever, dyspnea, and anosmia/ageusia, that demonstrated higher prevalence among infected patients, while coryza and sore throat were significant to Controls. In the COVID-19 group, 43.8% of cases presented mild symptoms and were redirected to homecare; for hospitalized patients, 28.1% received noninvasive oxygen support, and 6.3% required invasive mechanical ventilation. Overall, 3 patients from the COVID-19 group (8.3%) deceased in the hospital. Significant differences (p-value < 0.05) were observed among groups regarding pre-existing comorbidities such as diabetes and hypertension, presenting higher prevalence on COVID-19 diagnosed group. Patients’ characteristics are summarized in Table 1.

**Table 1.** Demographic characteristics of enrolled patients.

| Characteristics | COVID-19 = 64 | Control = 37 | *p*-value |
| --- | --- | --- | --- |
| Age in years, median (IQR) | 45 (22)<sup>a</sup> | 34 (21) | 0.005 |
| Female sex, N (%) | 38 (59.4) | 29 (78.4) | 0.052 |
| Severity, N (%) |  |  |  |
| *homecare* | 28 (43.8) | 36 (97.3) | < 0.001<sup>c</sup> |
| *hospitalization* | 36 (56.2) | 1 (2.7) |  |
| ≤ 10 days | 26 (72.2) | 0 | 0.293<sup>c</sup> |
| >10 days | 10 (27.8) | 1 (100) |  |
| Respiratory support, N (%) |  |  |  |
| *no oxygen received* | 42 (65.6) | 36 (97.3) | < 0.001<sup>c</sup> |
| *noninvasive oxygen received* | 18 (28.1) | 1 (2.7) |  |
| *invasive mechanical ventilation* | 4 (6.3) | 0 |  |
| Outcome death from hospitalized patients | 3 (8.3) | 0 | 1<sup>c</sup> |
| Onset of symptoms to enrolment in days, median (IQR) | 8 (6)<sup>b</sup> | 5.5 (4) | 0.031 |
| Symptoms |  |  |  |
| *fever* | 49 (76.6) | 18 (48.6) | 0.004 |
| *dry cough* | 47 (73.4) | 22 (59.5) | 0.146 |
| *wet cough* | 12 (18.8) | 8 (21.6) | 0.727 |
| *dyspnea* | 42 (65.6) | 14 (37.8) | 0.007 |
| *malaise* | 45 (70.3) | 32 (86.5) | 0.066 |
| *coryza* | 31 (48.4) | 31 (83.8) | < 0.001 |
| *sore throat* | 28 (43.8) | 25 (67.6) | 0.021 |
| *diarrhea* | 19 (29.7) | 9 (24.3) | 0.562 |
| *headache* | 39 (60.9) | 29 (78.4) | 0.072 |
| *anosmia / ageusia* | 32 (50.0) | 2 (5.4) | < 0.001 |
| Comorbidities, N (%) |  |  |  |
| *diabetes* | 14 (21.9) | 2 (5.4) | 0.029 |
| *hypertension* | 19 (29.7) | 4 (10.8) | 0.029 |
| *obesity* | 11 (17.2) | 1 (2.7) | 0.051<sup>c</sup> |
| *cardiomyopathy* | 6 (9.4) | 0 | 0.083<sup>c</sup> |
| *respiratory diseases* | 6 (9.4) | 4 (10.8) | 1<sup>c</sup> |
| *chronic renal diseases* | 1 (1.6) | 0 | 1<sup>c</sup> |
| *chronic hepatic diseases* | 0 | 0 | - |
| *HIV* | 0 | 0 | - |

<sup>a</sup>N = 63; <sup>b</sup>N = 61; <sup>c</sup> Fisher's Exact test.

### Metabolomic analysis of skin imprint

We employed an untargeted metabolomic strategy to identify potential biomarkers present in the COVID-19 patient’s skin that may contribute to disease diagnosis and pathophysiology assessment. Hence, a full mass spectra dataset in the *m/z* range of 150 to 1,700 was normalized, transformed, and analyzed using a supervised multivariate analysis, PLS-DA. Figure 2a shows the supervised PLS-DA plot discriminating the Control from COVID-19 group. From the PLS-DA-associated VIP score list, we were able to annotate 14 markers, from which 7 were found increased in COVID-19 patients and 7 decreased (elevated in controls). Detailed characterization of annotated markers is displayed in Supporting information Table S1. The putative annotations of enhanced markers (Figure 2a) belong to the class of primary fatty acid amides (PFAM) (palmitic amide, linoleamide, and oleamide), N-acylethanolamines (NAE) (palmitoleoyl-ethanolamide, POEA), N-acyl amino acids (N-palmitoyl threonine), and glycerolipids (a triacylglycerol (TG(24:0)) and a diacylglycerol (DG(40:10))). In contrast, decreased biomarkers consisted of phosphatidylserines (PS(34:0), PS(40:5), and PS(P-38:5)), dipeptides (cysteinyl-glutamine, valyl-arginine), and sterol lipids (cortisol and trihydroxyvitamin D3). Permutation tests (p-value < 0.001) were used to evaluate model predictability (data shown on Supporting information Figure S1).

**Figure 2.**
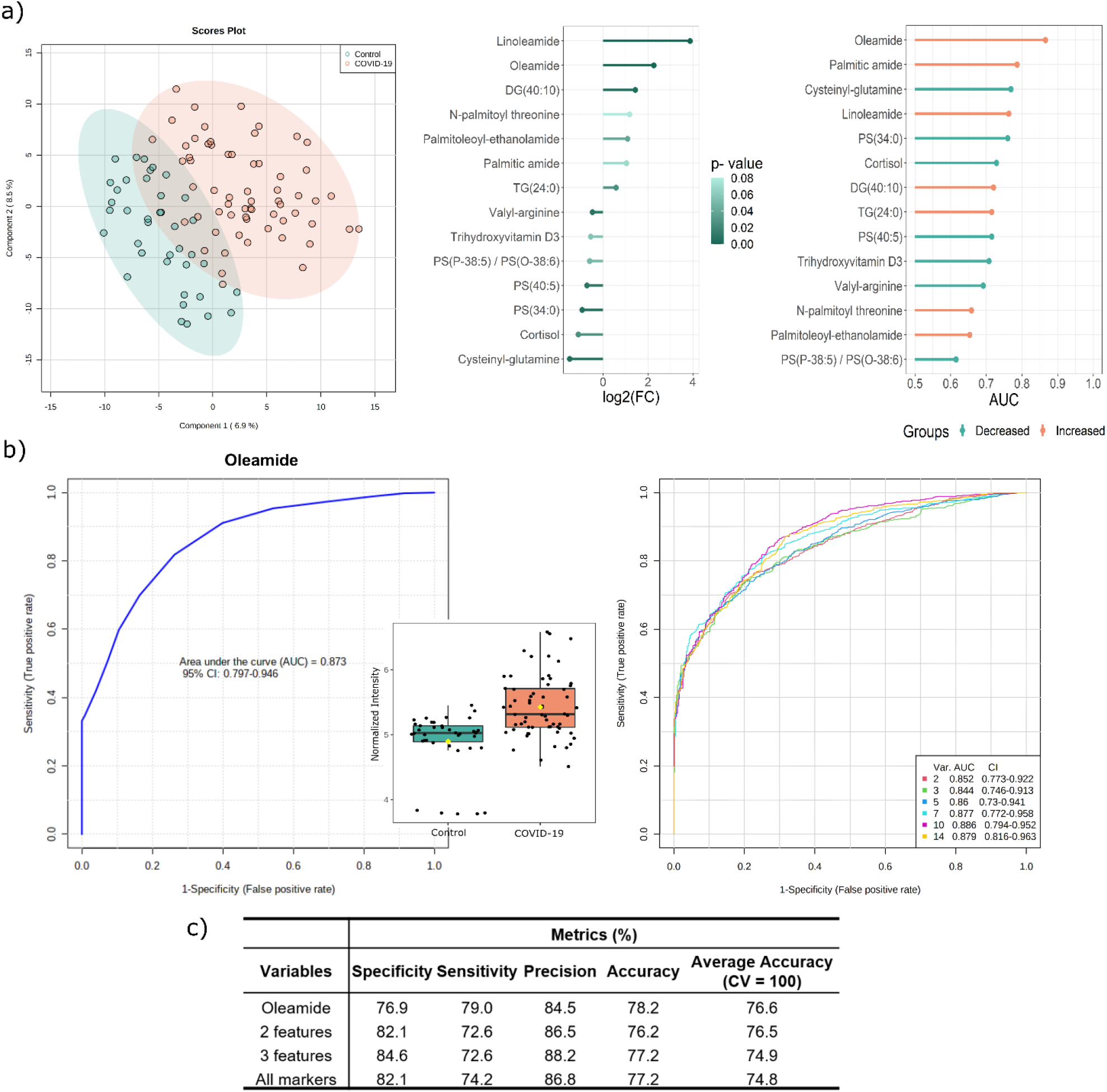
PLS-DA plot discriminated COVID-19 from Control group with listed annotated markers according to log_2_(FC) and p- value, and AUC (a). A ROC curve using Oleamide data, the most prominent marker ranked by AUC and a multivariate ROC curve was used to evaluate the influence of variable number (b) on model performance (c).

The importance of markers is ordered according to the fold-change of relative intensities from COVID-19 over the Control group, with associated p-value and area under curve (AUC) (Figure 2a). Oleamide, linoleamide, DG(40:10) for COVID-19 group and cysteinyl-glutamine and PS(34:0) for Control group presented significant alterations in the fold-change of their *m/z* feature intensities (p-value < 0.01). Even though linoleamide presented the highest difference in fold-change, oleamide exhibited the upmost AUC (0.873) (Figure 2b). Oleamide normalized intensities in COVID-19 and Control were used to project curve with a 95% of Confidence Interval from 0.797 to 0.946. The ROC model (Figure 2b) achieved a predicted accuracy of 76.6%, and calculated specificity and sensitivity of 79.0% and 76.9%, respectively; the model was validated by 1000 permutations (empiric p-value < 0.001). A multivariate receiver operating characteristic (ROC) was projected using 2, 3, 5, 7, 10 and 14 markers as variables (Figure 2b). The performance obtained with oleamide, 2 variables, 3 variables and all markers are exemplified in Figure 2c and detailed in Figure S2 e S3 available as supporting information. While the oleamide demonstrated the best sensitivity (79.0%), the highest specificity was obtained with 3 features (84.6%).

## DISCUSSION

COVID-19 has been pointed to as a multifactorial disease with multi-organ implications, especially due to the broad distribution of SARS-CoV-2 binding receptor, ACE2^14^. While the impairment of respiratory, nervous, cardiovascular, and gastrointestinal systems has been widely explored, skin involvement only gained notoriety with the increased reporting of skin lesions^14,15,17,22^. Xue et al. (2020) demonstrated that keratinocytes expressed high levels of ACE2, in particular differentiating keratinocytes and basal cells, while sweat gland cells presented moderate expression^15^. These findings suggested the importance of further investigation on the involvement of skin barrier during SARS-CoV-2 infection^15^. In addition to clinical and histopathological profiling of skin manifestations, metabolic alterations presented in normal skin of COVID-19 patients and the pathogenic mechanisms of the virus in this organ were little explored ^12,22,23^.

In a pilot study with 67 participants, Spick et al. (2021) demonstrated that the sebum lipidome of COVID-19 normal skin presented alterations in ceramides and glycerolipids when compared to controls. Particularly, a decrease in triacyglycerol levels was perceived, implying the importance of this biofluid for diagnostic testing ^12^. In our analysis, TG(24:0) and DG(40:10) were the representants of glycerolipids class and were decreased in the skin imprint of COVID-19 patients. Disrupted lipid metabolism is a common feature in viral infection, and particularly for COVID-19, altered levels of glycerolipids are associated with the presence and severity of SARS-CoV-2 infection on plasma and serum samples^4,5,8,9^. Among other lipid species, phosphatidylserines were cited as decreased metabolites in COVID-19^5,10,11,13^. Four previous studies reported reduced levels of PS in COVID-19 cases^5,11^, even when mild^10^, moderate and severe^13^ cases were compared to controls, on both blood-derived samples^5,10,13^ and naso-oropharyngeal swabs^11^, corroborating our findings. PS is a negatively-charged membrane phospholipid that may translocate from inner to outer cell membrane layer in early stages of viral infection^24,25^. It functions as a signal for macrophages cell phagocytoses and may also occur during platelet activation to contribute to the inflammatory and coagulopathy patterns observed in COVID-19^24,25^. Zaid et al. (2020) noticed a significant increase in platelet extracellular vesicles in patients with COVID-19 compared to healthy subjects, and a decrease in severe cases against mild manifestations. Besides, platelet EV exposing PS were significantly enhanced in non-severe symptoms, which may suggest its consumption and depletion in severe cases^25^.

In addition to other lipid classes, elevated PFAMs (palmitic amide, linoleamide, and oleamide) and NAE (palmitoleoyl-ethanolamide) species were interesting findings observed on COVID-19 group when compared to Controls. Fatty acid amides are part of the complex endocannabinoid system, and the biological mechanisms of several species are not fully elucidated^26^. However, in general, these bioactive lipids present relevant neuromodulatory and immune properties^26^. Even the cannabinoid receptor-inactive NAEs and PFAMs exert influence in the interaction of active endocannabinoids (e.g. anandamide) with receptors (CB1 and CB2). Due to site competition, cannabinoid receptor-inactive NAEs and PFAMs may delay the active molecules inactivation via FAAH (fatty acid amide hydrolase) and transport, keeping active forms available for longer time to interact with CB receptors^26,27^. While CB1 is primarily found in the brain and nervous tissues, CB2 is expressed in varied types of skin tissue cells (basal keratinocytes) and annexes (sweat gland duct, undifferentiated sebaceous cells, and undifferentiated infundibular hair follicle cells), in addition to immune cells (mast cells, monocytes/macrophages, B and T lymphocytes) showing a role in immunomodulation^27–29^. The immunosuppressive effect of CB2 receptors leads to a depletion of immune cell recruitment, macrophage polarization, modulation of T helper cells 1 and 2 activities, and proinflammatory cytokines and chemokines release^28,30,31^. Due to these immunosuppressive activities, the CB2 receptor has been pointed as a potential therapeutic target in COVID-19 management of cytokine storm^30,31^. However, the extent of immunomodulation should be carefully evaluated based on the SARS-CoV-2 infection timeline, since it may influence viral pathogenesis outcome^28,30^.

Oleamide, the most prominent marker found in our study is a bioactive PFAM metabolized by the FAAH enzyme^26^. Together with linoleamide, oleamide is known as a sleep-inducing agent, interacting with the GABA system^26^, Ca^2+^ transport modulation^26,32^, in addition to being a selective endogenous agonist of human CB1 receptor^33,34^. The exact mechanism by which oleamide promotes its biological function within the endocannabinoid system remains unclear, and so the allosteric regulation of CB1/CB2 has not been discarded^27,34^. Even though their bioactivity is associated with nervous and immune system stimulation, fatty acid amides have been detected in urine, plasma, sweat, and saliva^35^. Both linoleamide and oleamide were previously reported as markers of COVID-19 in nasal- and oropharyngeal samples when analyzed using paper-spray mass spectrometry technique^11^. In our study, the increase of oleamide alone in the COVID-19 patients’ skin achieved a good performance as a predictor of the disease, with specificity of 76.9% and sensitivity of 79.0%, reassuring its potential as a COVID-19 diagnosis biomarker.

Two other lipid metabolites, trihydroxyvitamin D3 and cortisol, were found in lower amounts on the skin imprint of COVID-19 patients. Several factors, such as stress, treatment, food supplementation, age, ethnicity, and latitude may influence sterols concentration, and therefore reliable levels of these substances to be considered as potential biomarkers for SARS-CoV-2 infection remain controversial and requires further data collection and validation^36–38^.

Altogether, the elected markers presented good specificity (82.1%) and sensitivity (74.2%) for COVID-19 screening, reinforcing the potential of alternative sampling methods for diagnostic tests. Even though markers were found correlated with some of the SARS-CoV-2 previously reported alterations, this cohort was enrolled in the first phase of Brazilian COVID-19 pandemic, and therefore, the investigation and validation of the results with a new sampling from the second wave may be appropriate. Most patients presented mild and moderate symptoms, with a low rate of invasive mechanical ventilation support and number of deceased patients. This demonstrates that the method was sensitive enough to discriminate COVID-19 from patients with mild flu-like symptoms. However, the incorporation of more severe cases would be beneficial for a tentative risk stratification as a future direction.

## CONCLUSION

The continued use of metabolomics as an approach for problem-solving in many fields of knowledge is providing insights of conditions and diseases at the biomolecular level. This information may serve as foundation, as well as validation of further studies and techniques. Our study aims to provide an overview on how the skin biochemistry is impacted by COVID-19 infection, demonstrating the main biomarkers present in this scenario, as well as the possible pathways affected by the biochemical disbalance, which may impact the endocannabinoid and immune system. Further investigation is required to pinpoint the actual causes and agents that are related to the widespread organ impairment in COVID-19 infection as an effort to provide better understanding and strategies to fight back this condition.

## Data Availability

Biomarkers elucidation data through mass spectrometry is available as supporting information
Pre-processed mass spectrometry data will be available at Zenodo following publication.
De-identified patient information will be made available from corresponding author upon request.

## ASSOCIATED CONTENT

Supporting Information

- Biomarker’s elucidation data through mass spectrometry (PDF).
- Pre-processed mass spectrometry data will be available at Zenodo following publication.
- De-identified patient information will be made available from corresponding author upon request.

## AUTHOR INFORMATION

Corresponding Author

* RRC -, phone +55 19 3521-9138

## AUTHOR CONTRIBUTIONS

RFS, JCN, TFD, AJB, ECS, MWPJ, LOR, ND, WJF, JD, and RRC were involved with study design. RFS, JCN, TFD, AJB, RS, FGM-B, ECS, AE, MWPJ, LOR, WJF, ND and LAS contributed to patient sample and data collection, clinical support, and network feasibility. JD, ENBB, GMS, ANO, DNO performed mass spectrometry experiments and data interpretation. JD and ANO performed data analysis and wrote the manuscript. DNO, RFS, JCN, MWPJ, ND, WJF and RRC revised the manuscript. RRC idealized the project and managed the research group. All authors approved the manuscript.

## ACKNOWLEDGMENTS

The authors would like to thank the network involved in sample collection, clinical data curation and diagnosis, in special to Paulínia Municipal Hospital and Sumaré State Hospital (CAAE 31049320.7.1001.5404) for providing samples to the initial steps of method development, and Thermo Scientific and LADETEC (UFRJ) for the technology support. This work was supported by São Paulo Research Foundation (FAPESP) [2019/05718-3 to JD, 2018/10052-1 to WJF, 2020/04705-2 to JCN and TFD], and Coordination for the Improvement of Higher Education Personnel (CAPES) [88887.513974/2020-00 to ANO].

## Competing Interests

The authors declare no conflict of interests.

## Notes

### Competing Interest Statement

The authors have declared no competing interest.

## REFERENCES

(1) Flaxman, S., Mishra, S., Gandy, A., Unwin, H. J. T., Mellan, T. A., Coupland, H., Whittaker, C., Zhu, H., Berah, T., Eaton, J. W., Monod, M., Perez-Guzman, P. N., Schmit, N., Cilloni, L., Ainslie, K. E. C., Baguelin, M., Boonyasiri, A., Boyd, O., Cattarino, L., Cooper, L. v., Cucunubá, Z., Cuomo-Dannenburg, G., Dighe, A., Djaafara, B., Dorigatti, I., van Elsland, S. L., FitzJohn, R. G., Gaythorpe, K. A. M., Geidelberg, L., Grassly, N. C., Green, W. D., Hallett, T., Hamlet, A., Hinsley, W., Jeffrey, B., Knock, E., Laydon, D. J., Nedjati-Gilani, G., Nouvellet, P., Parag, K. v., Siveroni, I., Thompson, H. A., Verity, R., Volz, E., Walters, C. E., Wang, H., Wang, Y., Watson, O. J., Winskill, P., Xi, X., Walker, P. G. T., Ghani, A. C., Donnelly, C. A., Riley, S., Vollmer, M. A. C., Ferguson, N. M., Okell, L. C., Bhatt, S. Estimating the Effects of Non-Pharmaceutical Interventions on COVID-19 in Europe. Nature 2020, 584 (7820), 257–261. https://doi.org/10.1038/s41586-020-2405-7.

(2) Jose, R. J., Manuel, A. COVID-19 Cytokine Storm: The Interplay between Inflammation and Coagulation. The Lancet Respiratory Medicine 2020, 8 (6), e46–e47. https://doi.org/10.1016/S2213-2600(20)30216-2.

(3) Caterino, M., Gelzo, M., Sol, S., Fedele, R., Annunziata, A., Calabrese, C., Fiorentino, G., D’Abbraccio, M., Dell’Isola, C., Fusco, F. M., Parrella, R., Fabbrocini, G., Gentile, I., Andolfo, I., Capasso, M., Costanzo, M., Daniele, A., Marchese, E., Polito, R., Russo, R., Missero, C., Ruoppolo, M., Castaldo, G. Dysregulation of Lipid Metabolism and Pathological Inflammation in Patients with COVID-19. Scientific Reports 2021, 11 (1), 1–10. https://doi.org/10.1038/s41598-021-82426-7.

(4) Shen, B., Yi, X., Sun, Y., Bi, X., Du, J., Zhang, C., Quan, S., Zhang, F., Sun, R., Qian, L., Ge, W., Liu, W., Liang, S., Chen, H., Zhang, Y., Li, J., Xu, J., He, Z., Chen, B., Wang, J., Yan, H., Zheng, Y., Wang, D., Zhu, J., Kong, Z., Kang, Z., Liang, X., Ding, X., Ruan, G., Xiang, N., Cai, X., Gao, H., Li, L., Li, S., Xiao, Q., Lu, T., Zhu, Y., Liu, H., Chen, H., Guo, T. Proteomic and Metabolomic Characterization of COVID-19 Patient Sera. Cell 2020, 182 (1), 59–72. https://doi.org/10.1016/j.cell.2020.05.032.

(5) Delafiori, J., Navarro, L. C., Siciliano, R. F., de Melo, G. C., Busanello, E. N. B., Nicolau, J. C., Sales, G. M., de Oliveira, A. N., Val, F. F. A., de Oliveira, D. N., Eguti, A., dos Santos, L. A., Dalçóquio, T. F., Bertolin, A. J., Abreu-Netto, R. L., Salsoso, R., Baía-Da-Silva, D., Marcondes-Braga, F. G., Sampaio, V. S., Judice, C. C., Costa, F. T. M., Durán, N., Perroud, M. W., Sabino, E. C., Lacerda, M. V. G., Reis, L. O., Fávaro, W. J., Monteiro, W. M., Rocha, A. R., Catharino, R. R. Covid-19 Automated Diagnosis and Risk Assessment through Metabolomics and Machine Learning. Analytical Chemistry 2021, 93 (4), 2471–2479. https://doi.org/10.1021/acs.analchem.0c04497.

(6) Grassin-Delyle, S., Roquencourt, C., Moine, P., Saffroy, G., Carn, S., Heming, N., Fleuriet, J., Salvator, H., Naline, E., Couderc, L. J., Devillier, P., Thévenot, E. A., Annane, D. Metabolomics of Exhaled Breath in Critically Ill COVID-19 Patients: A Pilot Study. EBioMedicine 2021, 63, 103154–undefined. https://doi.org/10.1016/j.ebiom.2020.103154.

(7) Costa Dos Santos Junior, G., Pereira, C. M., Kelly Da Silva Fidalgo, T., Valente, A. P. Saliva NMR-Based Metabolomics in the War against COVID-19. Analytical Chemistry 2020, 92 (24), 15688–15692. https://doi.org/10.1021/acs.analchem.0c04679.

(8) Su, Y., Chen, D., Yuan, D., Lausted, C., Choi, J., Dai, C. L., Voillet, V., Duvvuri, V. R., Scherler, K., Troisch, P., Baloni, P., Qin, G., Smith, B., Kornilov, S. A., Rostomily, C., Xu, A., Li, J., Dong, S., Rothchild, A., Zhou, J., Murray, K., Edmark, R., Hong, S., Heath, J. E., Earls, J., Zhang, R., Xie, J., Li, S., Roper, R., Jones, L., Zhou, Y., Rowen, L., Liu, R., Mackay, S., O’Mahony, D. S., Dale, C. R., Wallick, J. A., Algren, H. A., Zager, M. A., Wei, W., Price, N. D., Huang, S., Subramanian, N., Wang, K., Magis, A. T., Hadlock, J. J., Hood, L.; Aderem, A., Bluestone, J. A., Lanier, L. L., Greenberg, P. D., Gottardo, R., Davis, M. M., Goldman, J. D., Heath, J. R. Multi-Omics Resolves a Sharp Disease-State Shift between Mild and Moderate COVID-19. Cell 2020, 183 (6), 1479–1495. https://doi.org/10.1016/j.cell.2020.10.037.

(9) Wu, D., Shu, T., Yang, X., Song, J.-X., Zhang, M., Yao, C., Liu, W., Huang, M., Yu, Y., Yang, Q., Zhu, T., Xu, J., Mu, J., Wang, Y., Wang, H., Tang, T., Ren, Y., Wu, Y., Lin, S.-H., Qiu, Y., Zhang, D. Y., Shang, Y., Zhou, X. Plasma Metabolomic and Lipidomic Alterations Associated with COVID-19. National Science Review 2020, 7 (7), 1157–1168. https://doi.org/10.1093/nsr/nwaa086.

(10) Song, J. W., Lam, S. M., Fan, X., Cao, W. J., Wang, S. Y., Tian, H., Chua, G. H., Zhang, C., Meng, F. P., Xu, Z., Fu, J. L., Huang, L., Xia, P., Yang, T., Zhang, S., Li, B., Jiang, T. J., Wang, R., Wang, Z., Shi, M., Zhang, J. Y., Wang, F. S., Shui, G. Omics-Driven Systems Interrogation of Metabolic Dysregulation in COVID-19 Pathogenesis. Cell Metabolism 2020, 32 (2), 188–202. https://doi.org/10.1016/j.cmet.2020.06.016.

(11) de Silva, I. W., Nayek, S., Singh, V., Reddy, J., Granger, J. K., Verbeck, G. F. Paper Spray Mass Spectrometry Utilizing Teslin® Substrate for Rapid Detection of Lipid Metabolite Changes during COVID-19 Infection. Analyst 2020, 145 (17), 5725–5732. https://doi.org/10.1039/d0an01074j.

(12) Spick, M., Longman, K., Frampas, C., Lewis, H., Costa, C., Walters, D. D., Stewart, A., Wilde, M., Greener, D., Evetts, G., Trivedi, D., Barran, P., Pitt, A., Bailey, M. Changes to the Sebum Lipidome upon COVID-19 Infection Observed via Rapid Sampling from the Skin. EClinicalMedicine 2021, 33, 100786–undefined. https://doi.org/10.1016/j.eclinm.2021.100786.

(13) Schwarz, B., Sharma, L., Roberts, L., Peng, X., Bermejo, S., Leighton, I., Casanovas-Massana, A., Minasyan, M., Farhadian, S., Ko, A. I., dela Cruz, C. S., Bosio, C. M. Cutting Edge: Severe SARS-CoV-2 Infection in Humans Is Defined by a Shift in the Serum Lipidome, Resulting in Dysregulation of Eicosanoid Immune Mediators. The Journal of Immunology 2021, 206 (2), 329– 334. https://doi.org/10.4049/jimmunol.2001025.

(14) Garg, S., Garg, M., Prabhakar, N., Malhotra, P., Agarwal, R. Unraveling the Mystery of Covid-19 Cytokine Storm: From Skin to Organ Systems. Dermatologic Therapy 2020, 33 (6), e13859–undefined. https://doi.org/10.1111/dth.13859.

(15) Xue, X., Mi, Z., Wang, Z., Pang, Z., Liu, H., Zhang, F. High Expression of ACE2 on Keratinocytes Reveals Skin as a Potential Target for SARS-CoV-2. Journal of Investigative Dermatology 2021, 141 (1), 206–209. https://doi.org/10.1016/j.jid.2020.05.087.

(16) Guarneri, C., Rullo, E. V., Pavone, P., Berretta, M., Ceccarelli, M., Natale, A., Nunnari, G. Silent COVID-19: What Your Skin Can Reveal. The Lancet Infectious Diseases 2021, 21 (1), 24–25. https://doi.org/10.1016/S1473-3099(20)30402-3.

(17) Genovese, G., Moltrasio, C., Berti, E., Marzano, A. V. Skin Manifestations Associated with COVID-19: Current Knowledge and Future Perspectives. Dermatology 2021, 237 (1), 1–12. https://doi.org/10.1159/000512932.

(18) Lima, E. D. O., de Macedo, C. S., Esteves, C. Z., de Oliveira, D. N., Pessolani, M. C. V., Nery, J. A. D. C., Sarno, E. N., Catharino, R. R. Skin Imprinting in Silica Plates: A Potential Diagnostic Methodology for Leprosy Using High-Resolution Mass Spectrometry. Analytical Chemistry 2015, 87 (7), 3585–3592. https://doi.org/10.1021/acs.analchem.5b00097.

(19) Esteves, C. Z., Dias, L. de A., Lima, E. de O., Oliveira, D. N. de; Odir Rodrigues Melo, C. F., Delafiori, J., Souza Gomez, C.C., Ribeiro, J. D., Ribeiro, A. F., Levy, C. E., Catharino, R. R. Skin Biomarkers for Cystic Fibrosis: A Potential Non-Invasive Approach for Patient Screening. Frontiers in Pediatrics 2018, 5, 290–undefined. https://doi.org/10.3389/fped.2017.00290.

(20) Corman, V. M., Landt, O., Kaiser, M., Molenkamp, R., Meijer, A., Chu, D. K. W., Bleicker, T., Brünink, S., Schneider, J., Schmidt, M. L., Mulders, D. G. J. C., Haagmans, B. L., van der Veer, B., van den Brink, S., Wijsman, L., Goderski, G., Romette, J. L., Ellis, J., Zambon, M., Peiris, M., Goossens, H., Reusken, C., Koopmans, M. P. G., Drosten, C. Detection of 2019 Novel Coronavirus (2019-NCoV) by Real-Time RT-PCR. Eurosurveillance 2020, 25 (3), 2000045–undefined. https://doi.org/10.2807/1560-7917.ES.2020.25.3.2000045.

(21) Chong, J., Wishart, D. S., Xia, J. Using MetaboAnalyst 4.0 for Comprehensive and Integrative Metabolomics Data Analysis. Current Protocols in Bioinformatics 2019, 68 (1), e86–undefined. https://doi.org/10.1002/cpbi.86.

(22) Recalcati, S. Cutaneous Manifestations in COVID-19: A First Perspective. Journal of the European Academy of Dermatology and Venereology 2020, 34 (5), e212–e213. https://doi.org/10.1111/jdv.16387.

(23) Novak, N., Peng, W., Naegeli, M. C., Galvan, C., Kolm-Djamei, I., Brüggen, C., Cabanillas, B., Schmid-Grendelmeier, P., Catala, A. SARS-CoV-2, COVID-19, Skin and Immunology – What Do We Know so Far? Allergy: European Journal of Allergy and Clinical Immunology 2020, 76 (3), 698–713. https://doi.org/10.1111/all.14498.

(24) Argañaraz, G. A., Palmeira, J. da F., Argañaraz, E. R. Phosphatidylserine inside out: A Possible Underlying Mechanism in the Inflammation and Coagulation Abnormalities of COVID-19. Cell Communication and Signaling 2020, 18 (1), 1–10. https://doi.org/10.1186/s12964-020-00687-7.

(25) Zaid, Y., Puhm, F., Allaeys, I., Naya, A., Oudghiri, M., Khalki, L., Limami, Y., Zaid, N., Sadki, K., ben El Haj, R., Mahir, W., Belayachi, L., Belefquih, B., Benouda, A., Cheikh, A., Langlois, M. A., Cherrah, Y., Flamand, L., Guessous, F., Boilard, E. Platelets Can Associate with SARS - CoV-2 RNA and Are Hyperactivated in COVID-19. Circulation Research 2020, 127 (11), 1404– 1418. https://doi.org/10.1161/CIRCRESAHA.120.317703.

(26) Hiley, C. R., Hoi, P. M. Oleamide: A Fatty Acid Amide Signaling Molecule in the Cardiovascular System? Cardiovascular Drug Reviews 2007, 25 (1), 46–60. https://doi.org/10.1111/j.1527-3466.2007.00004.x.

(27) Fonseca, B. M., Costa, M. A., Almada, M., Correia-Da-Silva, G., Teixeira, N. A. Endogenous Cannabinoids Revisited: A Biochemistry Perspective. Prostaglandins and Other Lipid Mediators 2013, 102, 13–30. https://doi.org/10.1016/j.prostaglandins.2013.02.002.

(28) Sipe, J. C., Arbour, N., Gerber, A., Beutler, E. Reduced Endocannabinoid Immune Modulation by a Common Cannabinoid 2 (CB2) Receptor Gene Polymorphism: Possible Risk for Autoimmune Disorders. Journal of Leukocyte Biology 2005, 78 (1), 231–238. https://doi.org/10.1189/jlb.0205111.

(29) Ständer, S., Schmelz, M., Metze, D., Luger, T., Rukwied, R. Distribution of Cannabinoid Receptor 1 (CB1) and 2 (CB2) on Sensory Nerve Fibers and Adnexal Structures in Human Skin. Journal of Dermatological Science 2005, 38 (3), 177–188. https://doi.org/10.1016/j.jdermsci.2005.01.007.

(30) Nagoor Meeran, M. F., Sharma, C., Goyal, S. N., Kumar, S., Ojha, S. CB2 Receptor-Selective Agonists as Candidates for Targeting Infection, Inflammation, and Immunity in SARS-CoV-2 Infections. Drug Development Research 2020, 82 (1), 7–11. https://doi.org/10.1002/ddr.21752.

(31) Rossi, F., Tortora, C., Argenziano, M., di Paola, A., Punzo, F. Cannabinoid Receptor Type 2: A Possible Target in SARS-CoV-2 (CoV-19) Infection? International Journal of Molecular Sciences 2020, 21 (11), 3809–undefined. https://doi.org/10.3390/ijms21113809.

(32) Lo, Y. K., Tang, K. Y., Chang, W. N., Lu, C. H., Cheng, J. S., Lee, K. C., Chou, K. J., Liu, C. P., Chen, W. C., Su, W., Law, Y. P., Jan, C. R. Effect of Oleamide on Ca2+ Signaling in Human Bladder Cancer Cells. Biochemical Pharmacology 2001, 62 (10), 1363–1369. https://doi.org/10.1016/S0006-2952(01)00772-9.

(33) Leggett, J. D., Aspley, S., Beckett, S. R. G., D’Antona, A. M., Kendall, D. A., Kendall, D. A. Oleamide Is a Selective Endogenous Agonist of Rat and Human CB 1 Cannabinoid Receptor s. British Journal of Pharmacology 2004, 141 (2), 253–262. https://doi.org/10.1038/sj.bjp.0705607.

(34) Hernández-Cervantes, R., Méndez-Díaz, M., Prospéro-García, Ó., Morales-Montor, J. Immunoregulatory Role of Cannabinoids during Infectious Disease. NeuroImmunoModulation 2018, 24 (4–5), 183–199. https://doi.org/10.1159/000481824.

(35) Castillo-Peinado, L. S., López-Bascón, M. A., Mena-Bravo, A., Luque de Castro, M. D., Priego-Capote, F. Determination of Primary Fatty Acid Amides in Different Biological Fluids by LC – MS/MS in MRM Mode with Synthetic Deuterated Standards: Influence of Biofluid Matrix on Sample Preparation. Talanta 2019, 193, 29–36. https://doi.org/10.1016/j.talanta.2018.09.088.

(36) Rhodes, J. M., Subramanian, S., Laird, E., Griffin, G., Kenny, R. A. Perspective: Vitamin D Deficiency and COVID-19 Severity – Plausibly Linked by Latitude, Ethnicity, Impacts on Cytokines, ACE2 and Thrombosis. Journal of Internal Medicine 2021, 289 (1), 97–115. https://doi.org/10.1111/joim.13149.

(37) Tan, T., Khoo, B., Mills, E. G., Phylactou, M., Patel, B., Eng, P. C., Thurston, L., Muzi, B., Meeran, K., Prevost, A. T., Comninos, A. N., Abbara, A., Dhillo, W. S. Association between High Serum Total Cortisol Concentrations and Mortality from COVID-19. The Lancet Diabetes and Endocrinology. 2020, pp 659–660. https://doi.org/10.1016/S2213-8587(20)30216-3.

(38) Slominski, R. M., Stefan, J., Athar, M., Holick, M. F., Jett en, A. M, Slominski, A., Raman, C.. T. COVID-19 and Vitamin D: A Lesson from the Skin. Experimental Dermatology 2020, 29 (9), 885–890. https://doi.org/10.1111/exd.14170.

