## Supporting information Table S1 for "Skin imprints to provide noninvasive metabolic profiling of COVID-19 patients"

### Table of Contents

Table S1. Proposed decreased and increased markers on skin imprint of COVID-19 patients…………………………………………………………………………………………..………..S3

Figure S1. Permutation tests for PLS-DA model evaluation…………..………………………………………S4

Figure S2. ROC curve additional data using oleamide to build the model……….……………………………S5

Figure S3. ROC curve additional data using all markers to build the model……….………………………….S6

**Table S1.** Proposed decreased and increased markers on skin imprint of COVID-19 patients.

| Class | Molecule | Exact *m/z* | Adduct | Metlin ID | Molecular Formula | Error (ppm) |
|---|---|---|---|---|---|---|
| Decreased | Cortisol | 385.1967 | $[M+Na]^+$ | 272 | $C_{21}H_{30}O_5$ | 4.67 |
| | Cysteinyl-glutamine | 250.0863 | $[M+H]^+$ | 85687 | $C_8H_{15}N_3O_4S$ | -2.80 |
| | PS(34:0)[a] | 802.4998 | $[M+K]^+$ | 78552 | $C_{40}H_{78}NO_{10}P$ | -0.37 |
| | PS(40:5)[a] | 860.5415 | $[M+Na]^+$ | 78435 | $C_{46}H_{80}NO_{10}P$ | -0.35 |
| | PS(P-38:5)[a] / PS(O-38:6)[a] | 816.5154 | $[M+Na]^+$ | 78790 / 78735 | $C_{44}H_{76}NO_9P$ | -0.49 |
| Increased | Trihydroxyvitamin D3 (calcitetrol) | 455.3129 | $[M+Na]^+$ | 57898 | $C_{27}H_{44}O_4$ | 0.66 |
| | Valyl-arginine | 296.1702 | $[M+Na]^+$ | 86013 | $C_{11}H_{23}N_5O_3$ | -3.04 |
| | DG(40:10)[a] | 661.4857 | $[M+H]^+$ | 58892 | $C_{43}H_{64}O_5$ | -4.54 |
| | TG(24:0)[a] | 471.3655 | $[M+H]^+$ | 62029 | $C_{27}H_{50}O_6$ | 5.30 |
| | Oleamide | 282.2792 | $[M+H]^+$ | 4115 | $C_{18}H_{35}NO$ | -0.35 |
| | Linoleamide | 280.2634 | $[M+H]^+$ | 43435 | $C_{18}H_{33}NO$ | 0.36 |
| | Palmitic amide | 278.2454 | $[M+Na]^+$ | 62905 | $C_{16}H_{33}NO$ | 0.00 |
| | Palmitoleoyl-ethanolamide | 320.2568 | $[M+Na]^+$ | 46564 | $C_{18}H_{35}NO_2$ | -2.50 |
| | N-palmitoyl threonine | 380.2773 | $[M+Na]^+$ | 75489 | $C_{20}H_{39}NO_4$ | -0.53 |

Metabolites considering structural and geometric isomers. [a] Carbon number: double bond. Abbreviations: DG – Diacylglycerol; PS – Phosphatidylserine; TG – Triacylglycerol.

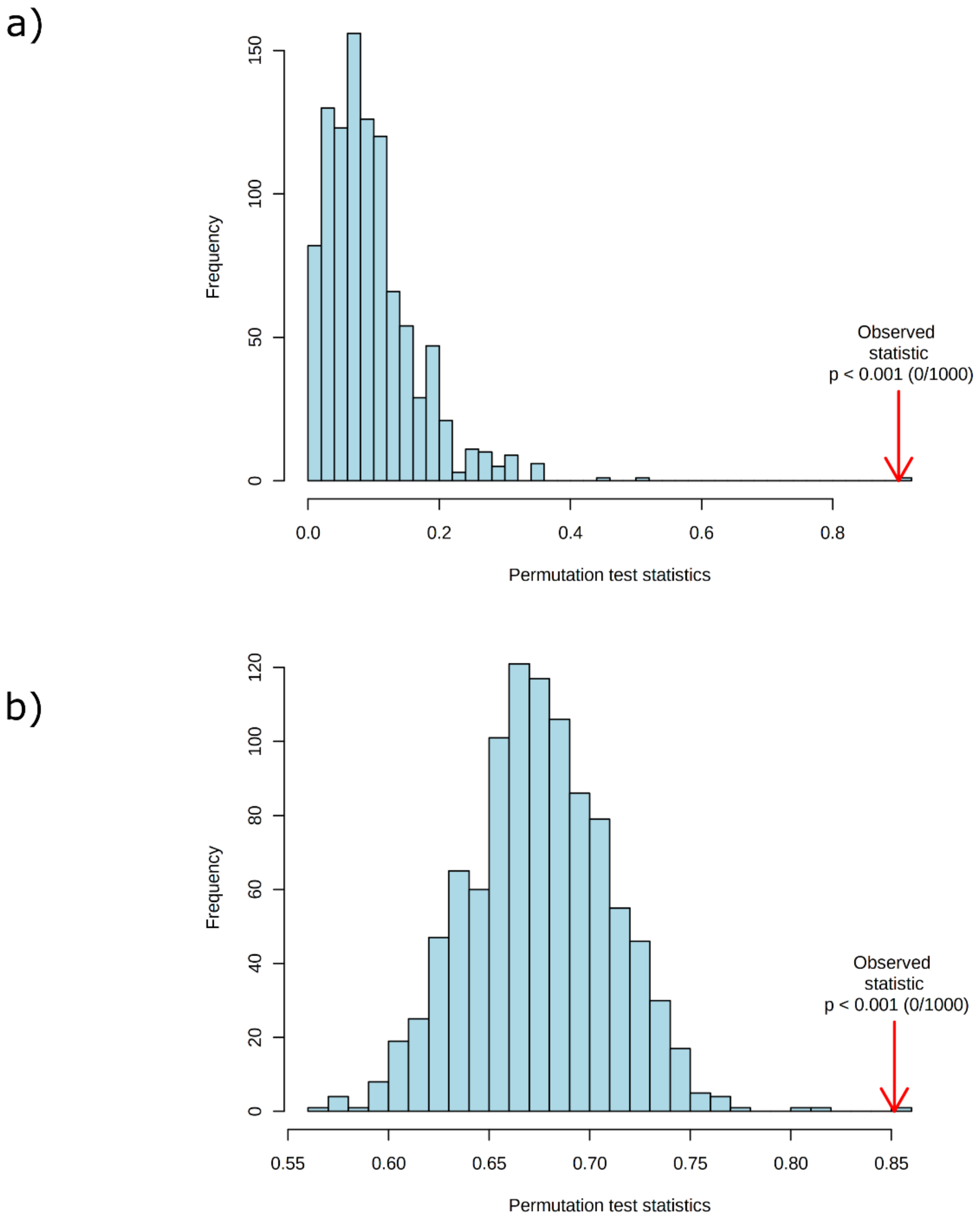


**Figure S1.** Permutation tests for PLS-DA model evaluation (n = 1000). (a) Separation distance. (b) Prediction accuracy during training.

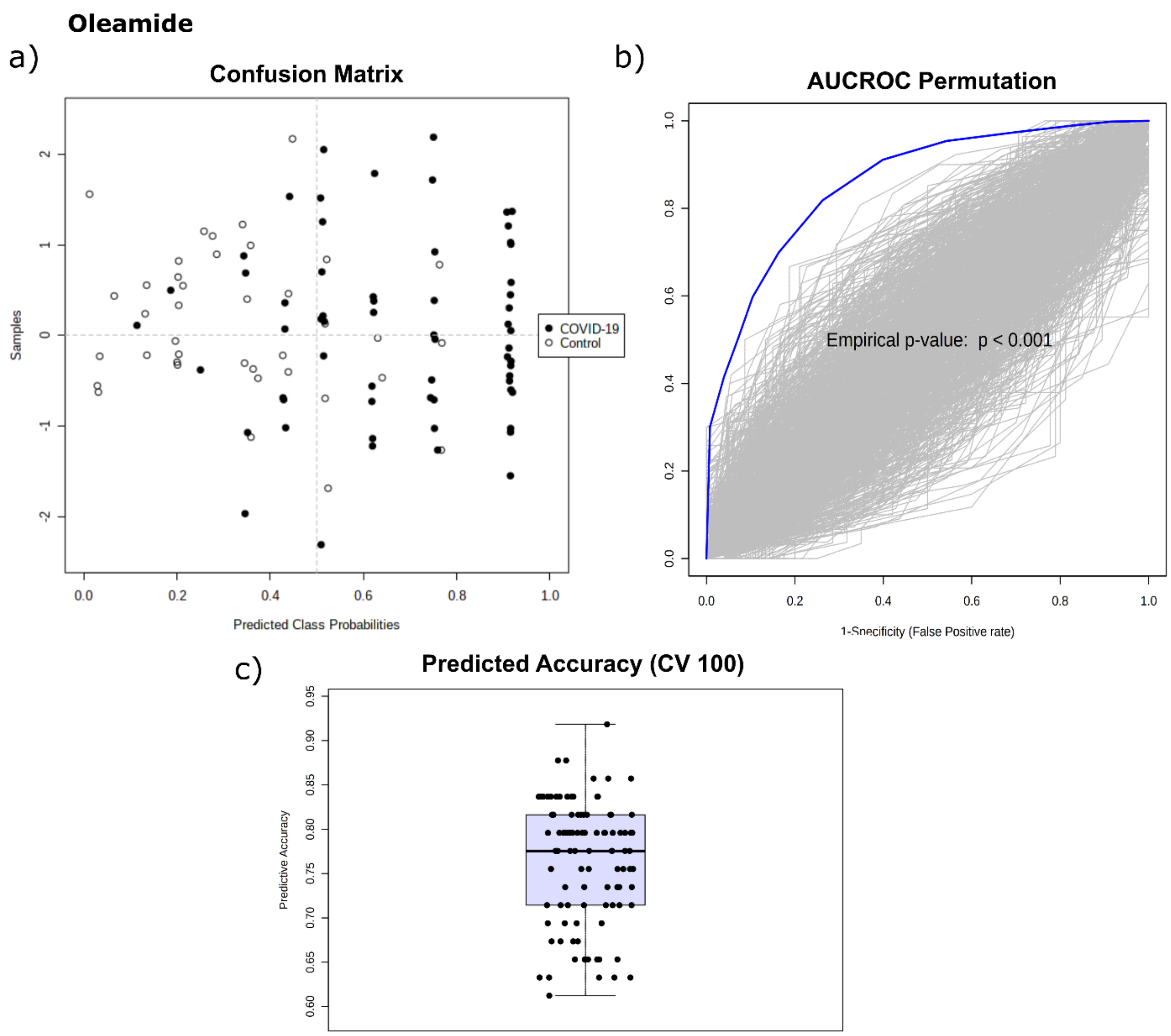


**Figure S2**. ROC curve additional data using oleamide to build the model. (a) Confusion matrix. (b) AUCROC permutation test (n = 1000) using Linear SVM algorithm. (c) Predicted accuracy during training (100 cross validations, CV).

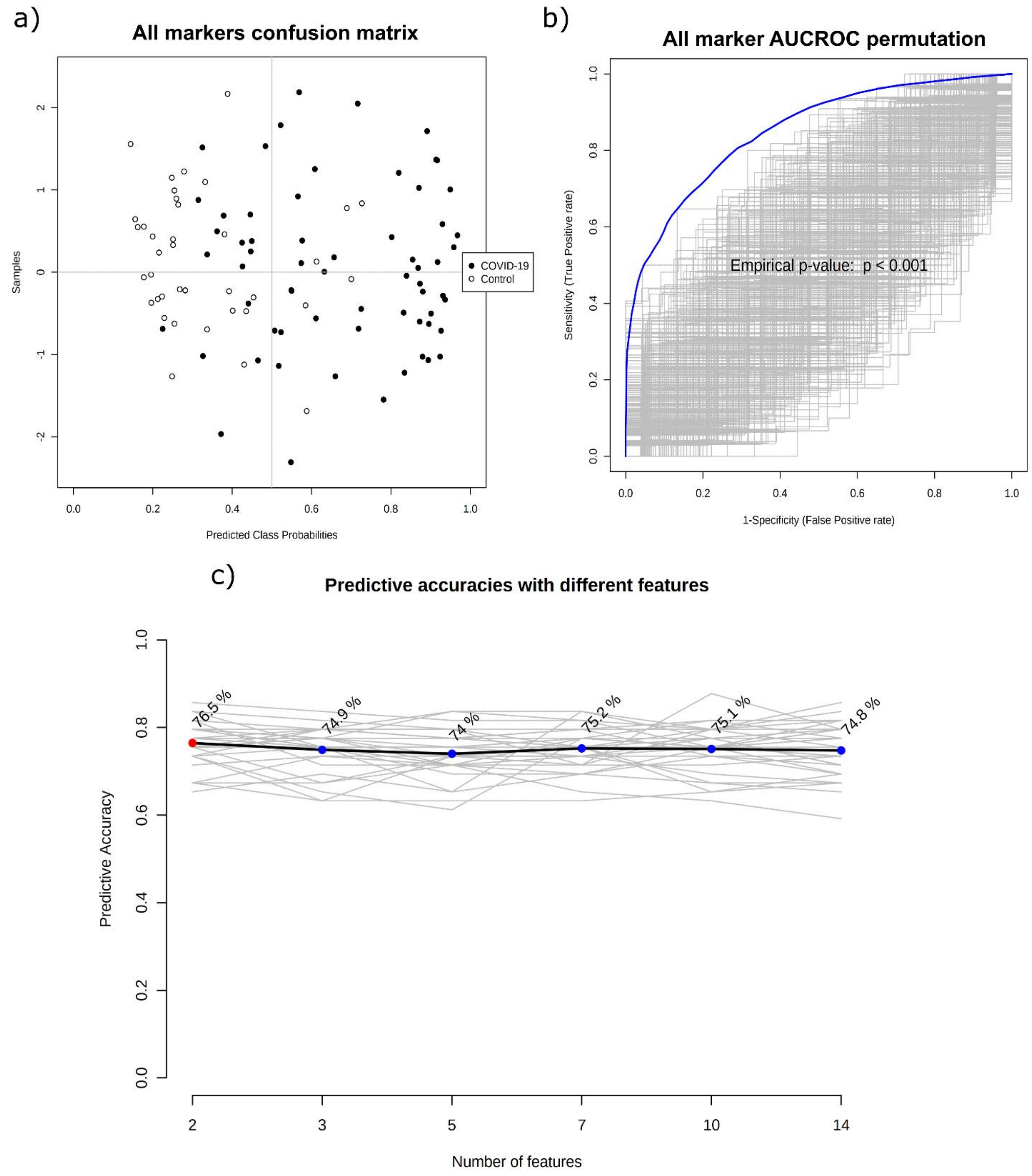


**Figure S3**. ROC curve additional data using all markers to build the model. (a) Confusion matrix. (b) AUCROC permutation test (n = 1000) using Linear SVM algorithm. (c) Predicted accuracies with different features.
